# United States’ political climates and the spread of SARS-2-COVID-19 during 2020

**DOI:** 10.1101/2022.05.16.22275162

**Authors:** Felicia Pratto, Andrew Cortopassi, Natasza Marrouch

**Affiliations:** Department of Psychological Sciences, University of Connecticut, Storrs, CT, USA; Universidade NOVA de Lisboa, Lisbon, Portugal

## Abstract

We tested whether the political climate in each U.S. state and Washington, DC determined the nature of the spread of COVID-19 cases and deaths in those polities during 2020. Political climate for each polity was indexed as a weighted average of the proportion of Republicans in legislatures in 2018 and the degree of public trust in both the White House and President Trump to handle COVID-19 in April, 2020. We found that polities higher on the political climate index had faster increases in per capita COVID-19 cases and deaths. Such Republican-trusting polities also had lower access to health care and less public engagement in prevention behavior, both of which mediated the influence of political climate on COVID- 19 cases and deaths. Further, the relationship between incidence of COVID-19 cases and deaths was weaker in more Republican-trusting polities. Political climate can be seen as contributing to more cases and deaths due to lower access to health care and to lower public adherence to public health guidelines in polities led by Republicans and which trusted the Trump White House to handle the pandemic.

## Introduction

History shows that the course of new communicable diseases can be influenced by the political climate in which they arrive. By “political climate,” we mean the socio-political agendas, ideologies, and practices that predominate in a populace during a particular place and time. The 1918 strain of influenza emerged in a political climate that prioritized US morale for the war effort. In fact, no national official acknowledged influenza’s dangers in words nor in policies, and local public health officials provided little or misleading information and minimized the terrible and mounting deaths people witnessed, touting *fear* of influenza as a greater threat than influenza itself. This “war morale” political climate guaranteed that the flu strain that originated in Kansas would be spread globally by moving infected troops to kill many millions. [1] In 1981, the HIV-AIDS epidemic emerged in the United States during a conservative rebound in which President Ronald Reagan had cut budgets at the Centers for Disease Control (CDC) and Food and Drug Administration; these budget cuts not only stymied prevention efforts for AIDS [2] but also fomented mistrust in federal agencies [3]. The devaluation of federal agencies and of the populations most impacted by AIDS ensured that the Reagan administration responded to AIDS with fundamental neglect, which contributed to its epidemic spread [2]. Thus, we see that the fear and uncertainty that dangerous new infectious disease brings can be multiplied, along with disease, when political climates prioritize other political agendas than conveying accurate information and protecting public health.

The present research examines how political climate influenced the sequelae of COVID-SARS-2 disease (COVID-19) in the United States prior to the possibility of immunization. COVID-19 is caused by a zoonotic virus, novel to humans, that is highly contagious via respiration from asymptomatic people [4]. Despite having the globe’s most extensive health care system, WHO data show that U.S. fatalities from COVID-19 exceed those of all other nations.[5] To understand whether political climate could have contributed to the spread of COVID-19 in the US and consequent deaths, we consider the political messages about the disease, which leaders the public trusted, and access to health care infrastructure. The present study tested how states’ political climates influenced the spread of COVID-19 in the U.S. and fatalities during 2020 when no vaccinations were available. Further, we tested whether two potential mediators of political climate -- access to health care, and public norms for disease-preventative behavior -- were proximate causes responsible for the effects of political climate.

### Republican-trusting political climate

We focus on the political climate created by the Republican Party because that party dominated all three branches of the federal government and most states during the period of our study. For over 50 years, Republican ideology has touted mistrust of and hostility towards government social programs. For example, President Reagan told people that help from the government should terrify them [3]. Republican strategist and lobbyist Grover Norquist said that his goal was to shrink government small enough that he could “drag it into the bathroom and drown it in the bathtub” [6]. While running for president and in office, Donald T. Trump fostered suspicion of government by promulgating the fiction that there is a “deep state” of government bureaucrats in charge whom no one else controls [7]. Many scholars analyze the Republicans’ messages touting the incompetence and malfeasance of government as an ideological narrative aimed at protecting the status quo and reversing the progress of the 20^th^ century civil rights movements by reducing support for anti-racist and other egalitarian government policies [8, 9]. By contrasting himself with the “fake news” media, “corrupt” Democrats, the “deep state,” etc., Donald Trump implies he is singularly trustworthy [10].

One recent Republican policy agenda has been to restrict access to health care. Unlike many developed nations, the U.S. has no national health insurance or sick leave policy. Instead, health insurance may be an employment benefit, and the US government offers public coverage only for the very poor (Medicaid) and subsidized insurance for elderly people (Medicare). In the 1990s and 2000s, the increasing costs of health care and growing unemployment led a number of Republicans, including Gov. Mitt Romney of Massachusetts, to advocate for states creating health insurance exchanges to provide regulated markets for individuals to buy health insurance [11]. However, when the Affordable Care Act (ACA), a national health insurance plan modelled on Gov. Romney’s state program, passed a Democrat-controlled Congress under a Democratic President (Obama) in 2010, it received no Republican votes. When Republicans gained controlled of the Congress in 2017, they made scores of unsuccessful attempts to repeal the ACA, President Trump having campaigned promising to “repeal and replace” the ACA [12]. Republican leaders in Congress and the White House conveyed, at best, misgivings about government health policy.

Though the ACA remains in place, Republicans have been more successful in restricting access to health care at the state level. Of the 15 states whose state governments refused to expand Medicaid under the ACA before 2020, all had a majority of at least 62% Republicans holding seats in the state legislature in 2018. In comparison, of the 36 polities (states and DC) that did expand Medicare, only 8 had 62% or more Republicans in their state legislature.^1^ Medicare expansion influences how many people have health insurance and therefore are likely to receive health care. In 2017, states that expanded Medicare coverage averaged 10.9% with no health insurance, whereas states that had expanded Medicare coverage averaged 6.7% uninsured.^2^ Hence, ideologically, mistrust of government, and policy-wise, restricting access to health care, are two COVID-relevant aspects of the political climate Republicans have fomented.

### Political face of COVID-19

Mistrust in government programs also characterized Republican responses to COVID-19. At a campaign rally in February, 2020, President Trump claimed that Democrats had politicized COVID-19, with the implication that their cautions or policy prescriptions should be disregarded [13]. In fact, President Trump sought to minimize the seriousness of COVID-19 and opposed shutting down public facilities and workplaces because he viewed a potential COVID-19-related economic crisis as a threat to his re-election [14]. In numerous public acts and practices, the White House linked defiance of public health practices with political identities. At the press conference on April 3^rd^, 2020 when the CDC first recommended that the public wear cloth masks to prevent airborne virus transmission, President Trump emphasized that doing so was optional and said he would not wear one. Estimates of the number of people who followed suit and then died of COVID-19 between that press conference and July 21, 2020, when President Trump recommended mask-wearing, range from approximately 4,000 to 12,000 [15]. White House protocols included no distancing or regular mask-wearing, and the White House hosted at least one super-spreader event [16].Trump did not wear a mask when returning to his residence after having been hospitalized for COVID-19 in October, 2020 [16], and his tweets that minimized his own case of COVID-19 persuaded some members of the public that COVID-19 is a hoax or not serious. [17]

The White House left COVID-19 strategies to the states, where political party control was strongly linked to COVID-prevention measures: Only 6 of 32 states that locked down in March, 2020 were controlled by Republicans in the executive and legislative branches; of the 18 states that did not lock down then, 14 were Republican-controlled, a reliable difference^3^ [18]. Republican governors were slowest to impose stay-at-home measures in the crucial early days of the pandemic [19]. From the White House down, undermining and refusing to follow public health recommendations to prevent COVID-19 spread became symbolically linked with Republican leaders and citizens, that is, politicized [19].

Such politicization of public health messages about COVID had the effect of dividing the US public concerning the threat of COVID-19 and measures to stop its spread [20]. Two US national convenience samples of MTurk workers and a 3000-person representative national survey in March 2020 found that self-identified Republicans and conservatives viewed COVID-19 as less of a threat to themselves and as being more hyped by the media than did Democrats, Independents, and liberals [21, 22, 23]. Concomittantly, internet searches about COVID-19 and unemployment in late February through March, 2020 were far less frequent in US counties that had higher vote share for Trump in the 2016 Presidential election than for other counties [27], suggesting less concern about COVID-19 in Trump- favoring polities. National random samples of the US conducted in April and June, 2020 showed that nearly all Democrats understood that individual behavior would influence the spread of COVID-19, whereas only 20% of Republicans acknowledged this [24]. Nationally-demographically representative samples found that those who identify as conservative (which is more typical of Republicans) assessed COVID-19 as less risky compared to moderates and liberals [25], and adhered more rarely to COVID-prevention behaviors such as mask-wearing [26]. On the whole, political messaging and political identities in the public divided the country as to whether COVID-19 presented a substantial threat and as to whether preventative-behaviors and preventative-policies should be followed.

Given COVID-19’s high level of contagion, people’s risk level is influenced by their local political climate because political climate is associated with how much people adhere to public health recommendations for preventing COVID-19 transmission. A national observational study in March-April, 2020 found that the level that states’ residents restricted their mobility (i.e., “sheltering-in-place” to limit viral exposure) was negatively related to the proportion of those who identified with Republicans in 2018 in those states [27]. Similarly, an observational study from late January into July 2020 found that mobility restriction was negatively related to the proportion of county votes cast for Republicans [28]. During the first resurgence of COVID-19 during June, 2020, states with Republican governors were more likely to have increases in case counts than states with Democratic governors [29]. Such findings suggest that more Republican-adhering political climates pose more risk of COVID-19 because in them, people adhere less to COVID-19 prevention behaviors such as avoiding crowds.

Considering political climate at the state level, which is where most COVID-19 public health measures were initiated [30], the present study examined the trajectories of COVID-19 cases and deaths over the days of 2020 in the US. Given that health care access has been a partisan issue, and given evidence that political party identification is associated with adherence to COVID-19 preventative behaviors [27, 29, 30, 32, 33, 34], we also tested whether indicators of access to health care and public adherence to COVID- 19 prevention practices could be responsible for the expected political climate effects. Thus, our study is unique in surveying the entire nation over the course of 2020 and in considering whether access to health care and public adherence to COVID-19 prevention practices mediated effects of general political climate.

### Predictions

Our pre-registered predictions, data, and analysis code can be found at https://osf.io/tqc45. IRB approval was not required because we used archival anonymous data.

US COVID-19 arrived in the US in more Democratic states on both coasts, but we expected that polities that favored Republican leaders would increase COVID-19 transmission, hence,

**Hypothesis 1:** Incidence of COVID-cases and deaths will begin higher per capita in polities lower on Republican-trusting political climate, but then will increase more strongly in polities that are higher on Republican-trusting political climate.

**Hypothesis 2**: Polities with more Republican-trusting political climates will show lower compliance with COVID prevention measures (e.g., wearing masks, avoiding crowds).

Due to Hypothesis 2,

**Hypothesis 3:** The effects of political climate predicted by Hypothesis 1 will be at least partly explained by polity-level differences in adherence to COVID-preventative behaviors.

Further, given that Republican-controlled states restricted access to health care, we propose,

**Hypothesis 4:** The effects of political climate predicted by Hypothesis 1 will be at least partly explained by polity-level differences in access to health care.

## Materials and methods

### Measures

The outcome measures were the number of COVID-19 cases (variable name “tot_cases”) and COVID-19 deaths (“tot_deaths”) per day within each U.S. State and Washington, DC for all available dates in 2020 (CDC, 2021) [31]. Our study therefore considered the time trajectories of COVID-19 cases and deaths for each polity.

State-level predictors included political climate, demographic controls (population size, population age 85 and older, percent of population who are People of Color [32], proportion of urban land mass), access to health care as measured by the number of federally funded clinics, patient visits, service sites per capita, percent of the state with no health insurance, and the number of Intensive Care Unit Beds per capita. Prior studies of part of 2020 show more COVID-19 cases and deaths for counties with more Blacks and which were more rural [32]. Table 1 lists demographic and health care access predictors by polity.

**Table 1.**
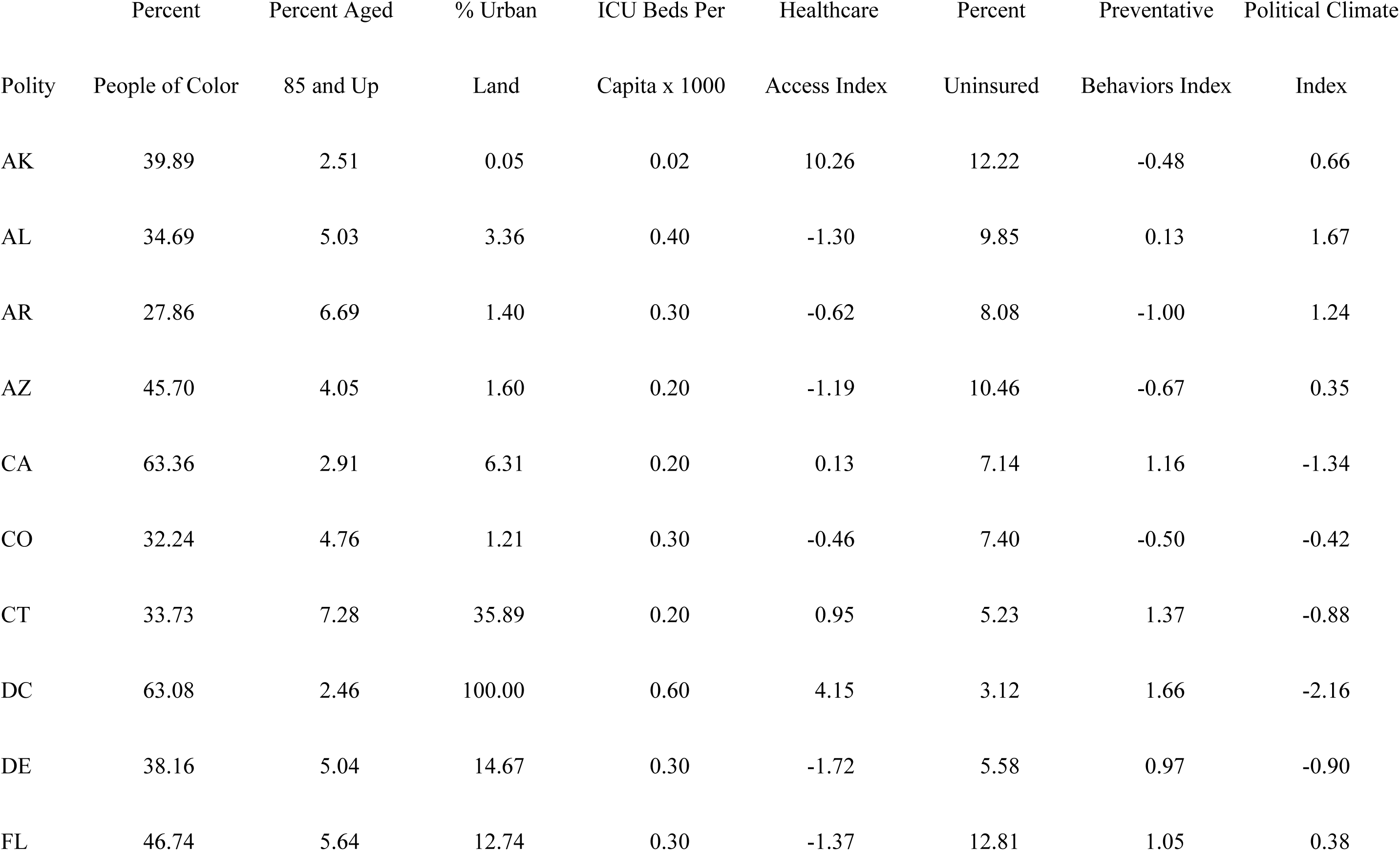
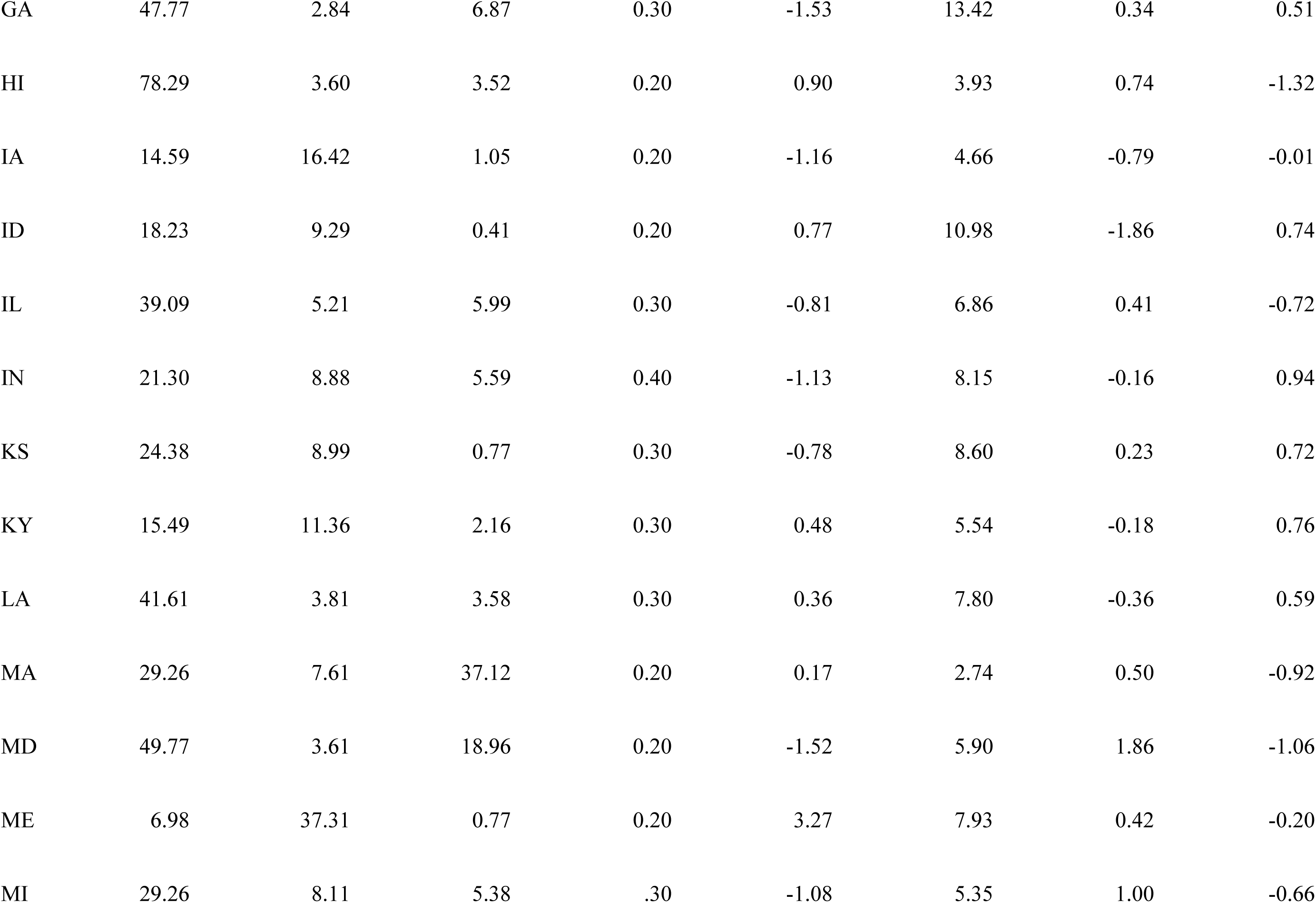
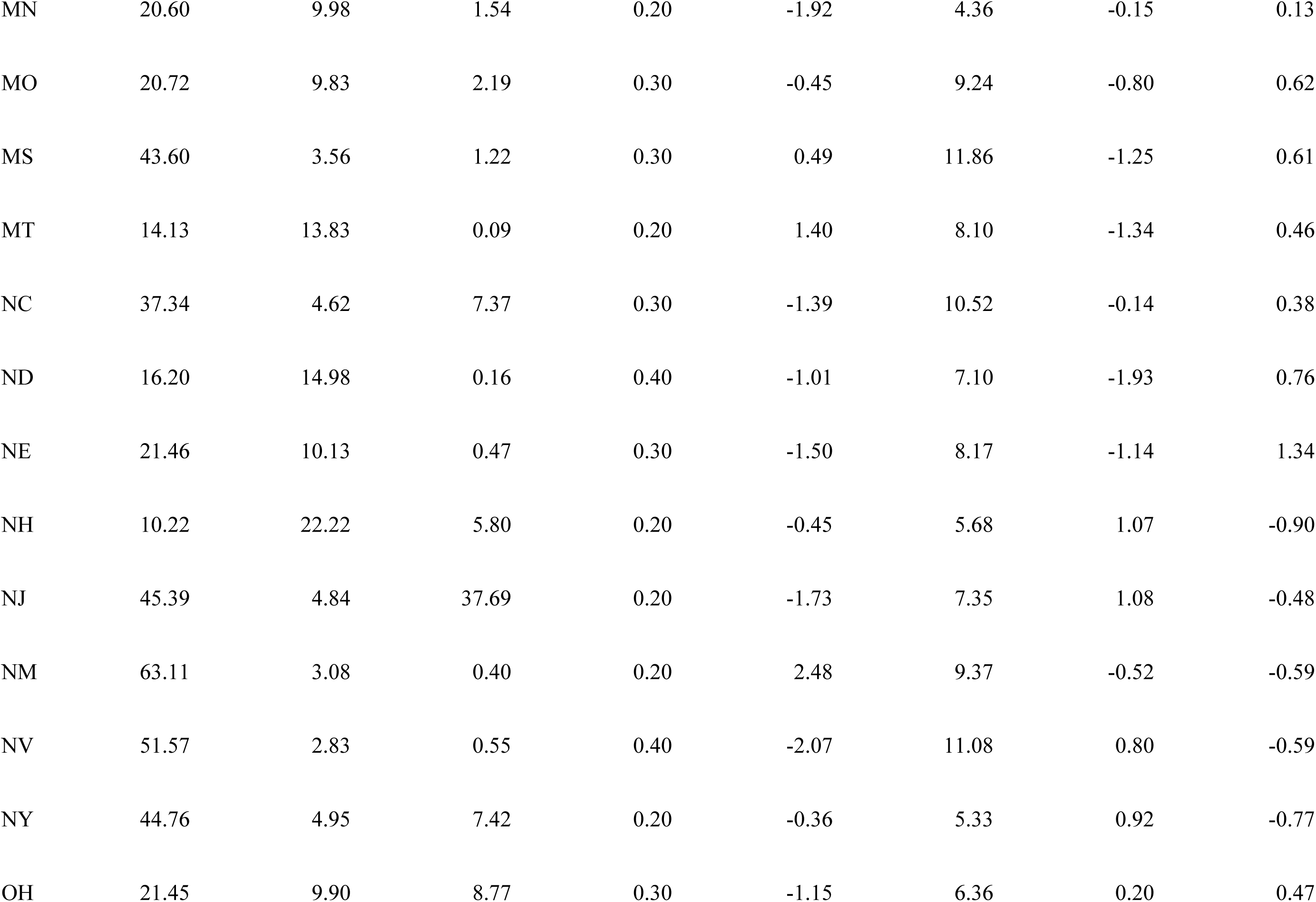
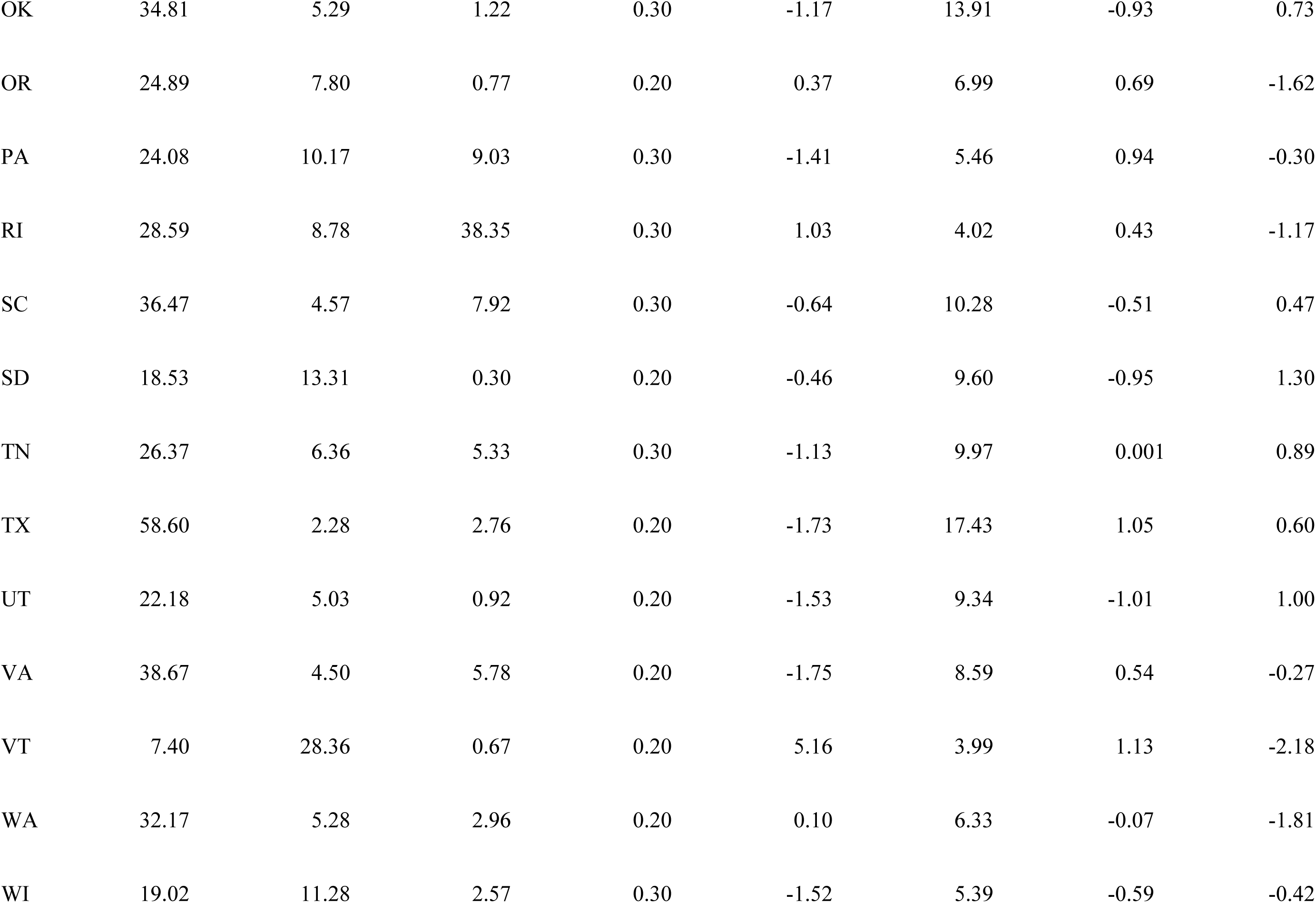
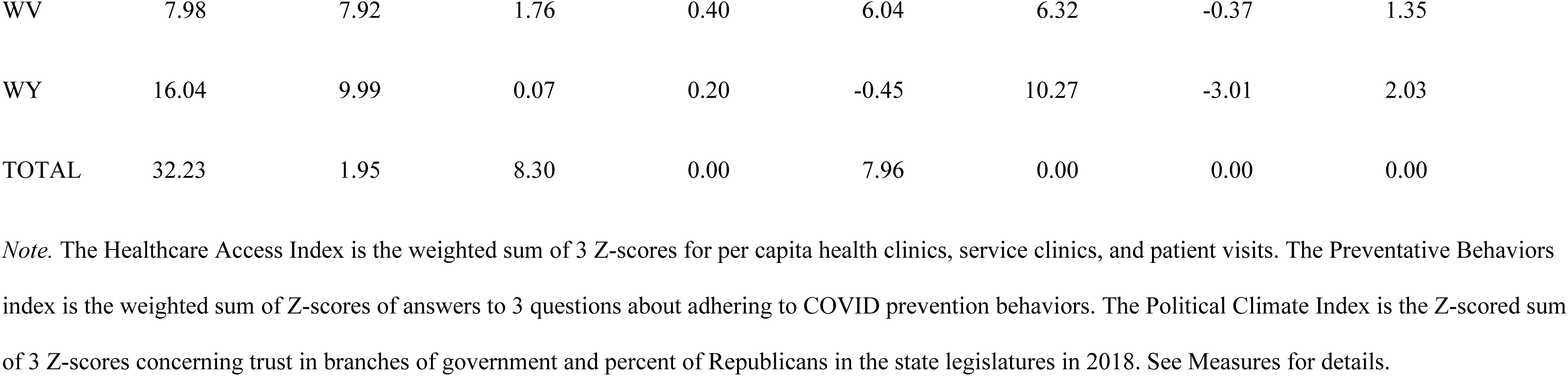
Demographic information, health access measures, and political climate index for each state.

| 179 |  | Percent | Percent Aged | % Urban | ICU Beds Per | Healthcare | Percent | Preventative | Political Climate |
| --- | --- | --- | --- | --- | --- | --- | --- | --- | --- |
| 180 | Polity | People of Color | 85 and Up | Land | Capita x 1000 | Access Index | Uninsured | Behaviors Index | Index |
| 181 | AK | 39.89 | 2.51 | 0.05 | 0.02 | 10.26 | 12.22 | -0.48 | 0.66 |
| 182 | AL | 34.69 | 5.03 | 3.36 | 0.40 | -1.30 | 9.85 | 0.13 | 1.67 |
| 183 | AR | 27.86 | 6.69 | 1.40 | 0.30 | -0.62 | 8.08 | -1.00 | 1.24 |
| 184 | AZ | 45.70 | 4.05 | 1.60 | 0.20 | -1.19 | 10.46 | -0.67 | 0.35 |
| 185 | CA | 63.36 | 2.91 | 6.31 | 0.20 | 0.13 | 7.14 | 1.16 | -1.34 |
| 186 | CO | 32.24 | 4.76 | 1.21 | 0.30 | -0.46 | 7.40 | -0.50 | -0.42 |
| 187 | CT | 33.73 | 7.28 | 35.89 | 0.20 | 0.95 | 5.23 | 1.37 | -0.88 |
| 188 | DC | 63.08 | 2.46 | 100.00 | 0.60 | 4.15 | 3.12 | 1.66 | -2.16 |
| 189 | DE | 38.16 | 5.04 | 14.67 | 0.30 | -1.72 | 5.58 | 0.97 | -0.90 |
| 190 | FL | 46.74 | 5.64 | 12.74 | 0.30 | -1.37 | 12.81 | 1.05 | 0.38 |
| 191 | GA | 47.77 | 2.84 | 6.87 | 0.30 | -1.53 | 13.42 | 0.34 | 0.51 |
| 192 | HI | 78.29 | 3.60 | 3.52 | 0.20 | 0.90 | 3.93 | 0.74 | -1.32 |
| 193 | IA | 14.59 | 16.42 | 1.05 | 0.20 | -1.16 | 4.66 | -0.79 | -0.01 |
| 194 | ID | 18.23 | 9.29 | 0.41 | 0.20 | 0.77 | 10.98 | -1.86 | 0.74 |
| 195 | IL | 39.09 | 5.21 | 5.99 | 0.30 | -0.81 | 6.86 | 0.41 | -0.72 |
| 196 | IN | 21.30 | 8.88 | 5.59 | 0.40 | -1.13 | 8.15 | -0.16 | 0.94 |
| 197 | KS | 24.38 | 8.99 | 0.77 | 0.30 | -0.78 | 8.60 | 0.23 | 0.72 |
| 198 | KY | 15.49 | 11.36 | 2.16 | 0.30 | 0.48 | 5.54 | -0.18 | 0.76 |
| 199 | LA | 41.61 | 3.81 | 3.58 | 0.30 | 0.36 | 7.80 | -0.36 | 0.59 |
| 200 | MA | 29.26 | 7.61 | 37.12 | 0.20 | 0.17 | 2.74 | 0.50 | -0.92 |
| 201 | MD | 49.77 | 3.61 | 18.96 | 0.20 | -1.52 | 5.90 | 1.86 | -1.06 |
| 202 | ME | 6.98 | 37.31 | 0.77 | 0.20 | 3.27 | 7.93 | 0.42 | -0.20 |
| 203 | MI | 29.26 | 8.11 | 5.38 | .30 | -1.08 | 5.35 | 1.00 | -0.66 |
| 204 | MN | 20.60 | 9.98 | 1.54 | 0.20 | -1.92 | 4.36 | -0.15 | 0.13 |
| 205 | MO | 20.72 | 9.83 | 2.19 | 0.30 | -0.45 | 9.24 | -0.80 | 0.62 |
| 206 | MS | 43.60 | 3.56 | 1.22 | 0.30 | 0.49 | 11.86 | -1.25 | 0.61 |
| 207 | MT | 14.13 | 13.83 | 0.09 | 0.20 | 1.40 | 8.10 | -1.34 | 0.46 |
| 208 | NC | 37.34 | 4.62 | 7.37 | 0.30 | -1.39 | 10.52 | -0.14 | 0.38 |
| 209 | ND | 16.20 | 14.98 | 0.16 | 0.40 | -1.01 | 7.10 | -1.93 | 0.76 |
| 210 | NE | 21.46 | 10.13 | 0.47 | 0.30 | -1.50 | 8.17 | -1.14 | 1.34 |
| 211 | NH | 10.22 | 22.22 | 5.80 | 0.20 | -0.45 | 5.68 | 1.07 | -0.90 |
| 212 | NJ | 45.39 | 4.84 | 37.69 | 0.20 | -1.73 | 7.35 | 1.08 | -0.48 |
| 213 | NM | 63.11 | 3.08 | 0.40 | 0.20 | 2.48 | 9.37 | -0.52 | -0.59 |
| 214 | NV | 51.57 | 2.83 | 0.55 | 0.40 | -2.07 | 11.08 | 0.80 | -0.59 |
| 215 | NY | 44.76 | 4.95 | 7.42 | 0.20 | -0.36 | 5.33 | 0.92 | -0.77 |
| 216 | OH | 21.45 | 9.90 | 8.77 | 0.30 | -1.15 | 6.36 | 0.20 | 0.47 |
| 217 | OK | 34.81 | 5.29 | 1.22 | 0.30 | -1.17 | 13.91 | -0.93 | 0.73 |
| 218 | OR | 24.89 | 7.80 | 0.77 | 0.20 | 0.37 | 6.99 | 0.69 | -1.62 |
| 219 | PA | 24.08 | 10.17 | 9.03 | 0.30 | -1.41 | 5.46 | 0.94 | -0.30 |
| 220 | RI | 28.59 | 8.78 | 38.35 | 0.30 | 1.03 | 4.02 | 0.43 | -1.17 |
| 221 | SC | 36.47 | 4.57 | 7.92 | 0.30 | -0.64 | 10.28 | -0.51 | 0.47 |
| 222 | SD | 18.53 | 13.31 | 0.30 | 0.20 | -0.46 | 9.60 | -0.95 | 1.30 |
| 223 | TN | 26.37 | 6.36 | 5.33 | 0.30 | -1.13 | 9.97 | 0.001 | 0.89 |
| 224 | TX | 58.60 | 2.28 | 2.76 | 0.20 | -1.73 | 17.43 | 1.05 | 0.60 |
| 225 | UT | 22.18 | 5.03 | 0.92 | 0.20 | -1.53 | 9.34 | -1.01 | 1.00 |
| 226 | VA | 38.67 | 4.50 | 5.78 | 0.20 | -1.75 | 8.59 | 0.54 | -0.27 |
| 227 | VT | 7.40 | 28.36 | 0.67 | 0.20 | 5.16 | 3.99 | 1.13 | -2.18 |
| 228 | WA | 32.17 | 5.28 | 2.96 | 0.20 | 0.10 | 6.33 | -0.07 | -1.81 |
| 229 | WI | 19.02 | 11.28 | 2.57 | 0.30 | -1.52 | 5.39 | -0.59 | -0.42 |
| 230 | WV | 7.98 | 7.92 | 1.76 | 0.40 | 6.04 | 6.32 | -0.37 | 1.35 |
| 231 | WY | 16.04 | 9.99 | 0.07 | 0.20 | -0.45 | 10.27 | -3.01 | 2.03 |
| 232 | TOTAL | 32.23 | 1.95 | 8.30 | 0.00 | 7.96 | 0.00 | 0.00 | 0.00 |

*Note.* The Healthcare Access Index is the weighted sum of 3 Z-scores for per capita health clinics, service clinics, and patient visits. The Preventative Behaviors index is the weighted sum of Z-scores of answers to 3 questions about adhering to COVID prevention behaviors. The Political Climate Index is the Z-scored sum of 3 Z-scores concerning trust in branches of government and percent of Republicans in the state legislatures in 2018. See Measures for details.

COVID-preventative behaviors were measured in a large national opinion poll conducted in April, 2020 [33] which asked, “In the past week, how closely did you follow the health recommendations listed below” and we used “wearing a face mask when outside your home, “avoiding contact with other people,” and “avoiding public or crowded places.” Following the opinion poll authors [33], we calculated a score for each state weighting 0 for the percent of people who selected “not at all closely,” 1 for the percent who selected “not very closely,” 2 for the percent who selected “somewhat closely” and 3 for the percent of people who selected “very closely.” The same opinion poll asked, “How much do you trust the following people and organizations to do the right thing to best handle the current coronavirus (COVID-19) outbreak?” and we used the answers to “The White House,” and “Donald Trump.” Each state’s score weighted the percent of people answering 0 for “not at all” 1 for “not too much,” 2 for “some” and 3 for “a lot.”

Using multiple measures to assess conceptual constructs is best practice [34, 35]. Analysis Step 1 was to test models of latent factors for political climate, for health care access, and for COVID-19 prevention behaviors. Confirmatory Factor Analysis (CFA) using the lavaan package in R [36] showed that the percentage of Republicans in the 2018 state legislature or DC city council, the trust the publics had in President Trump and in the White House to handle COVID-19 in April, 2020 formed a reliable political climate latent factor. Likewise, adherence to COVID-prevention behaviors (“wearing masks,” “avoiding crowds,” and “avoiding close contact with other people”) formed a reliable latent factor (see fit statistics in Table 2). CFA of the number of federally funded clinics per capita, health service centers per capita, patient visits per capita, the number of Intensive Care Unit (ICU) beds per capita, and the percent of people with no health insurance, showed that a factor that included the first three measures converged (see Table 2).

**Table 2.** Results of latent variable models for predictors of daily COVID-19 cases and deaths.

261 Latent Factor: Political climate
| 262 | Variable | estimate | SE | Z | p-value |
| --- | --- | --- | --- | --- | --- |
| 263 | Trust in Pres. Trump to handle COVID-19 | 1.00 |  |  |  |
| 264 | Percent of Republicans in legislature | .688 | .087 | 9.76 | < .001 |
| 265 | Trust in White House to handle COVID-19 | .845 | .108 | 6.34 | < .001 |

$\chi^2(1) = .05, p = .82$ . CFI = 1.00, TLI = 1.00. RMSEA = .00 [0, .225], SRMR = .031 Latent Factor: Access to Health Care
| 268 | Variable | estimate | SE | Z | p-value |
| --- | --- | --- | --- | --- | --- |
| 269 | Community Health sites per capita | 1.00 |  |  |  |
| 270 | Patient Center visits per capita | 0.662 | 0.113 | 5.87 | < .001 |
| 271 | Health Service Centers per capita | 0.836 | .091 | 9.18 | < .001 |

$\chi^2(1) = 0.07, p = .79$ . CFI = 1.00, TLI = 1.00, RMSEA = 0.00 [0, .239], SRMR = .035 Latent Factor: Endorsement of COVID-19 prevention behaviors
| 274 | Variable | estimate | SE | Z | p-value |
| --- | --- | --- | --- | --- | --- |
| 275 | Avoiding public places and crowds | 1.00 |  |  |  |
| 276 | Avoiding close contact with other people | 0.916 | .107 | 8.58 | < .001 |
| 277 | Wearing face masks in public | 0.618 | .124 | 5.00 | < .001 |

$\chi^2(1) = 0.25, p = .62$ . CFI = 1.00, TLI = 1.00. RMSEA = 0 [0, .29], SRMR = .066
*Note.* All variables were Z-scored before confirmatory factor analysis. N = 51. 90% CIs are shown for RMSEA in [].

Therefore, when considering whether health care access influenced COVID cases and COVID deaths, we used three separate and largely independent measures: the medical access factor score, the percent of uninsured, and per capita ICU beds. Table 3 shows correlations among all polity-level predictors.

**Table 3.** Correlations among polity-level variables.

| 281 |  | Medical access | % Uninsured | ICU Beds | Medicare | COVID-prevention |
| --- | --- | --- | --- | --- | --- | --- |
| 282 |  | index | in population | per capita | expanded | behavior index |
| 283 | Political climate index | -.15 | .53*** | -.04 | -.44*** | -.67*** |
| 284 | Medical access index |  | -.12 | -.33* | .24 | .00 |
| 285 | Percent uninsured people |  |  | .29* | -.64*** | -.32* |
| 286 | No. ICU Beds per capita |  |  |  | -.17 | .38** |
| 287 | State expanded Medicare under ACA |  |  |  |  | .32* |

*Note.* N = 51 cases (50 states plus DC). State expanded Medicare under ACA is coded 1 if it had by 2019 and 0 if it had not. \* $p < .05$ , \*\* $p < .01$ , \*\*\* $p < .001$ .
Indices were scores on the latent factors defined in Table 2.

### Analyses

Case and death counts for each polity began on the first “outbreak” day on which the polity had a case count of 40 or over. The data have a nested structure: day within polity. The most specific (level 1) data are each day’s COVID case and death count, which are nested within polity (group) at level 2.

Predictor variables such as political climate are associated with polity at level 2. Hypotheses 1, 3, and 4 concern how level 2 measures influence the time (level 1) patterns. To accurately interpret results from level 2 variables, one must first specify a correct model to explain the level 1 time effects [37]. We employed the lmer package in R [38] to test all multi-level models. Step 2 was to identify the best level 1 time model for each outcome variable, and test whether there remained variability in the slopes and intercepts at level 2. For nested models, we compared improvement in model fits using χ^2^. For non-nested models, we used the AIC and BIC and pseudo-R^2^ to identify the best-fitting model. To interpret findings, we compared the estimates to check whether a predictor reduced the effect of a correlated predictor or whether suppression effects occurred.

We tested several functions of COVID-19 cases and deaths (reciprocal, powers, ln, ln divided by polity population) and comparing multiple time periods (see Tables S2-S8).^4^ Given that population size influences case and death counts and that the natural log function described the start of the outbreak, we analyzed the natural log of cases and deaths per capita (times 10,000 for scaling) to test hypotheses. We initially included both polity-linked time metric (variable called “StateDay” in Supplementary materials) and day of 2020 (variable called “Day1” in Supplementary materials) and dropped day of 2020 when it was unreliable. StateDay was a random effect, and in interaction with predictor variables was a fixed effect.

Outbreaks began between days 65 and 85 of 2020. The best “base” time models are Model 6 in Table S4a for COVID-19 cases and Model 10 in Table S8 for COVID-19 deaths.

In Step 3, we tested for the influence of each demographic variable entered singly, and then in combination, by adding the demographic level 2 variables to the base time models. We trimmed unreliable effects (*ps* ≥ .0551) before adding another class of level 2 predictors.

In Step 4, we added hypothesized level 2 variables (i.e., political climate, health care access, and preventative behavior indices) and their interactions with time parameters to the base models.

For robustness, we tested whether the political climate index moderated base model time effects with and without demographic effects (Models 1 and 2 in Tables S12, S13 for cases, deaths, respectively). Though we report demographics when entered alone in their raw form, to increase model estimability, we Z-scored demographic variables when entering hypothesized (and Z-scored) level 2 variables in addition.

## Results

We first explain how COVID-19 cases and deaths changed over time. We then explain whether demographic variables influenced the time trajectories. Finally, we report the hypothesized effects of political climate and whether it appeared to be mediated, as predicted, by health care access and preventative behavior.

### Time models

Cases and deaths over time showed the typical natural log pattern, followed by an uptick towards the end of 2020 (see Fig 1 for COVID-19 deaths and Supplement Fig S4 for COVID-19 cases). We therefore modelled the outcome variables (which were the natural log of COVID-19 cases and deaths per 10,000 people for each day for each polity) using three linear slopes. In the models, each period has an intercept corresponding to the number of cases or deaths on the first day of the period (per polity per

**Fig 1.**
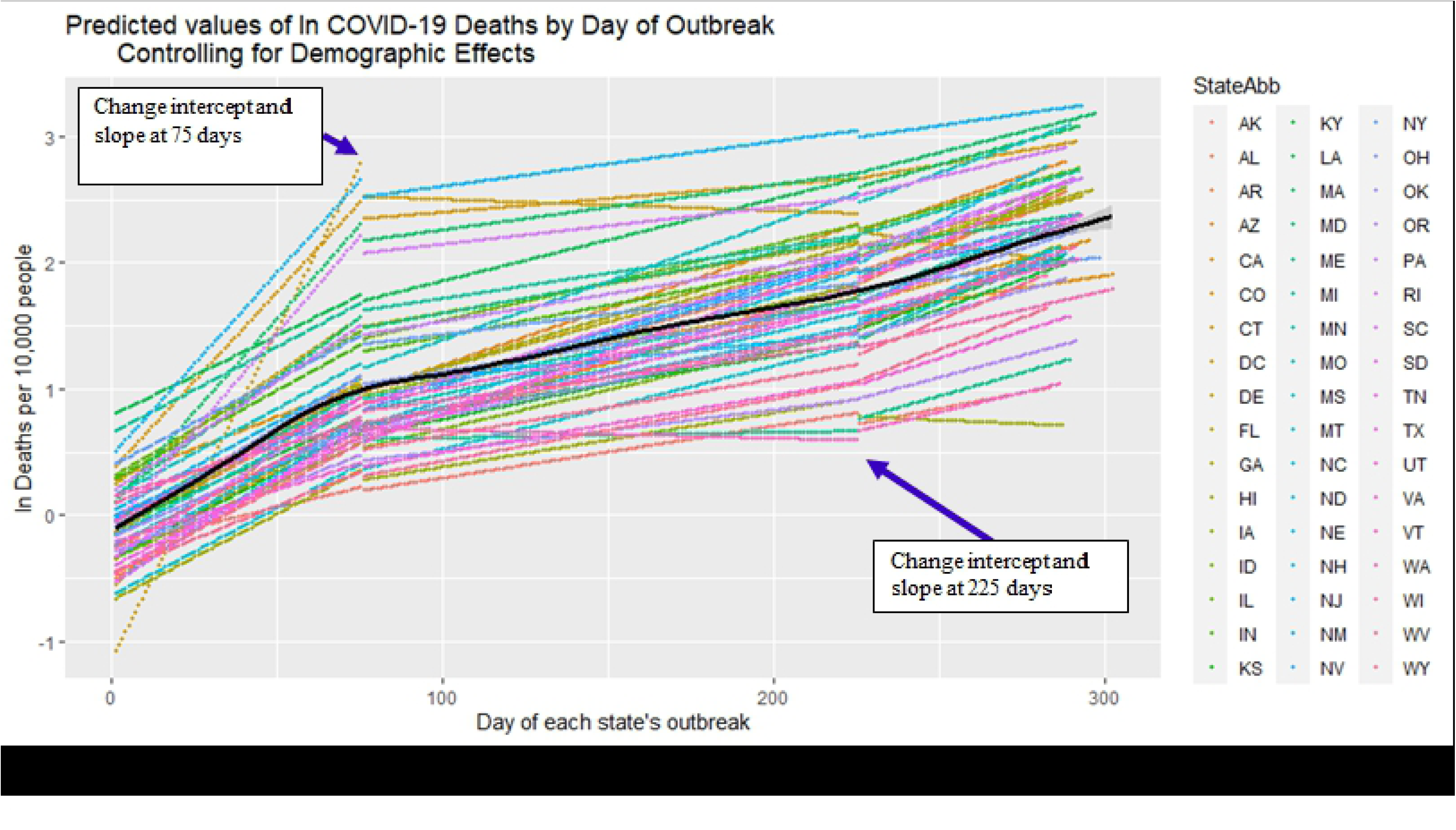
Ln COVID-19 deaths per day per 10,000 people. The black line represents the average across polities. Colors represent polities (abbreviation denoted as StateAbb). Arrows show where the model calculated a new intercept and slope at 75 days and 225 days, in addition to the intercept and slope at the first day of outbreak.

10,000 people) and a slope (percent daily change). Exponentiating terms, the base time model showed that on average there were 1.88 COVID-19 cases per day per polity per 10,000 people in the population (these aspects of rates are hereafter not stated) at the outset of the initial period with an increase of 6% per day through day 50; then on day 51 there averaged 19.49 COVID-19 cases with a 1% increase per day through day 150; then an intercept of 10.28 COVID-19 cases on day 151 with a 2% increase through the end of 2020 (see Model 6 in Table S4a).

For deaths, day of calendar year was a reliable effect; for each day later in the year the intercept reduced by 4%. For a polity with the average calendar day, the base time model had 0.90 COVID-19 deaths on the first day of the initial period with an increase of 2% per day through outbreak day 75, then an intercept of 1.82 deaths with a 1% daily increase through outbreak day 225, followed by an intercept of 1.28 deaths on outbreak day 226 and a 1.2% daily increase through the end of 2020 (Model 10 in Table S8).

Cases and deaths both showed substantial variability associated with polity (fixed effects pseudo- R^2^ was 84.38% for cases; 59.20% for deaths), justifying further models to test the influence of polity-level variables. Different demographic variables influenced COVID-19 cases and deaths in different periods (see Tables S9 & S10). The fact that demographic effects changed in time shows that the pattern of COVID-19 cases and deaths are not merely attributable to demographic factors.

### Political Climate

The political climate index was standardized (average = 0 and units are standard deviations).

Positive values indicate polities being above average on Republicans elected and on trusting in Trump and the White House to handle COVID-19; negative values indicate being below average on those measures (see Method and Table 2).

Table 4 shows the exponentiated intercepts and slopes for cases and deaths and how much they changed as a function of the political climate index in each of the three periods. As Fig 2 shows, cases increased more quickly in the initial period for the least Republican-trusting states (shown with shades of blue), but this pattern reversed directions in middle and final periods (see Sections 1 and 2 of Table 4). As Sections 5 and 6 of Table 4 show, the same pattern of higher initial increases in slopes for polities low on the political climate index, followed by higher slope increases for Republican-trusting polities, also occurred for COVID-19 deaths. These findings are consistent with Hypothesis 1.

**Fig 2.**
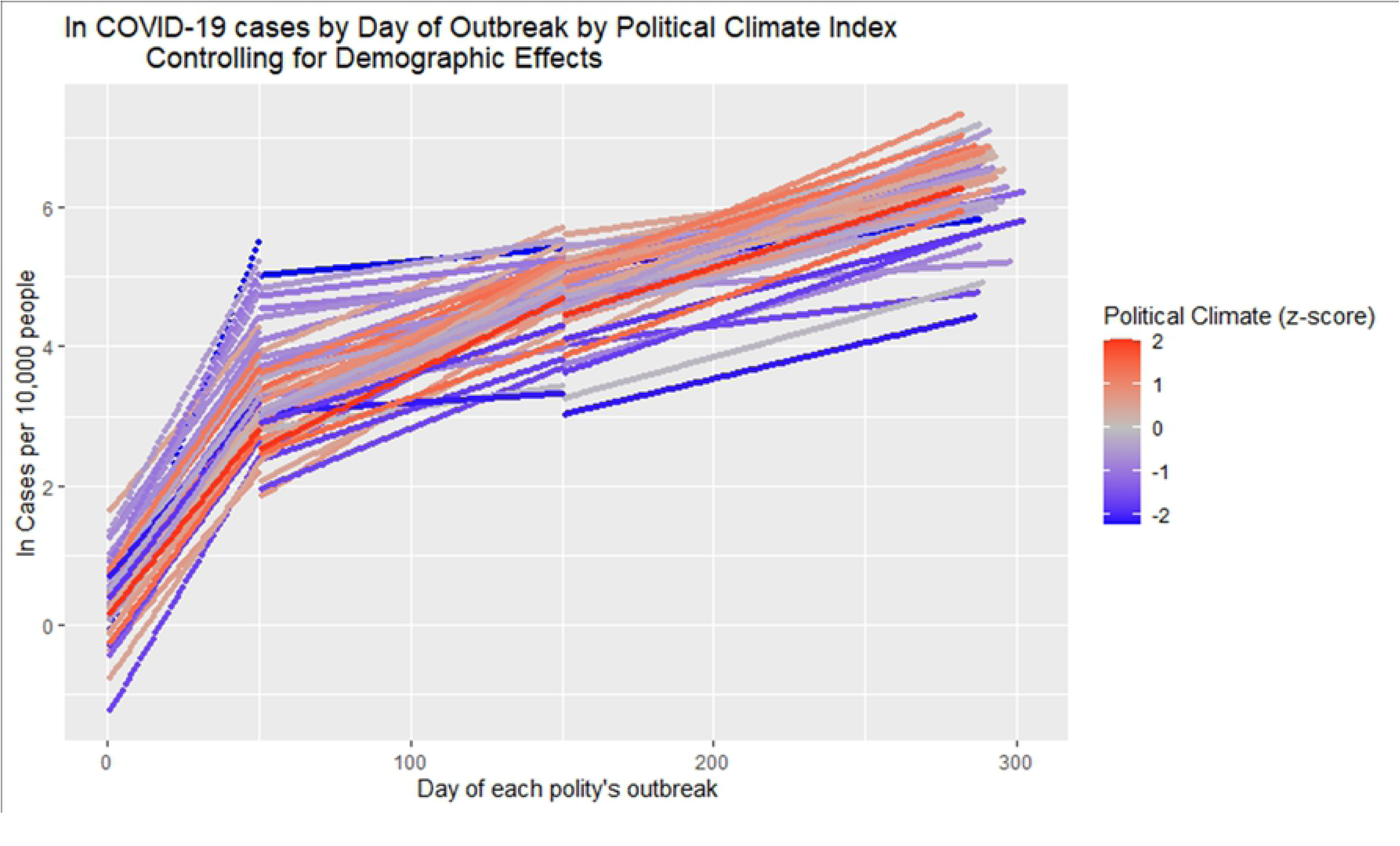
Model-predicted ln COVID-19 cases per 10,000 people as a function of day of polities’ outbreak and polities’ political climate index. (which is Z scored; ZPOLCLIMATEF). The model calculated an intercept and slope at Day 1, Day 51, and Day 151 of each polities’ outbreak.

**Table 4.**
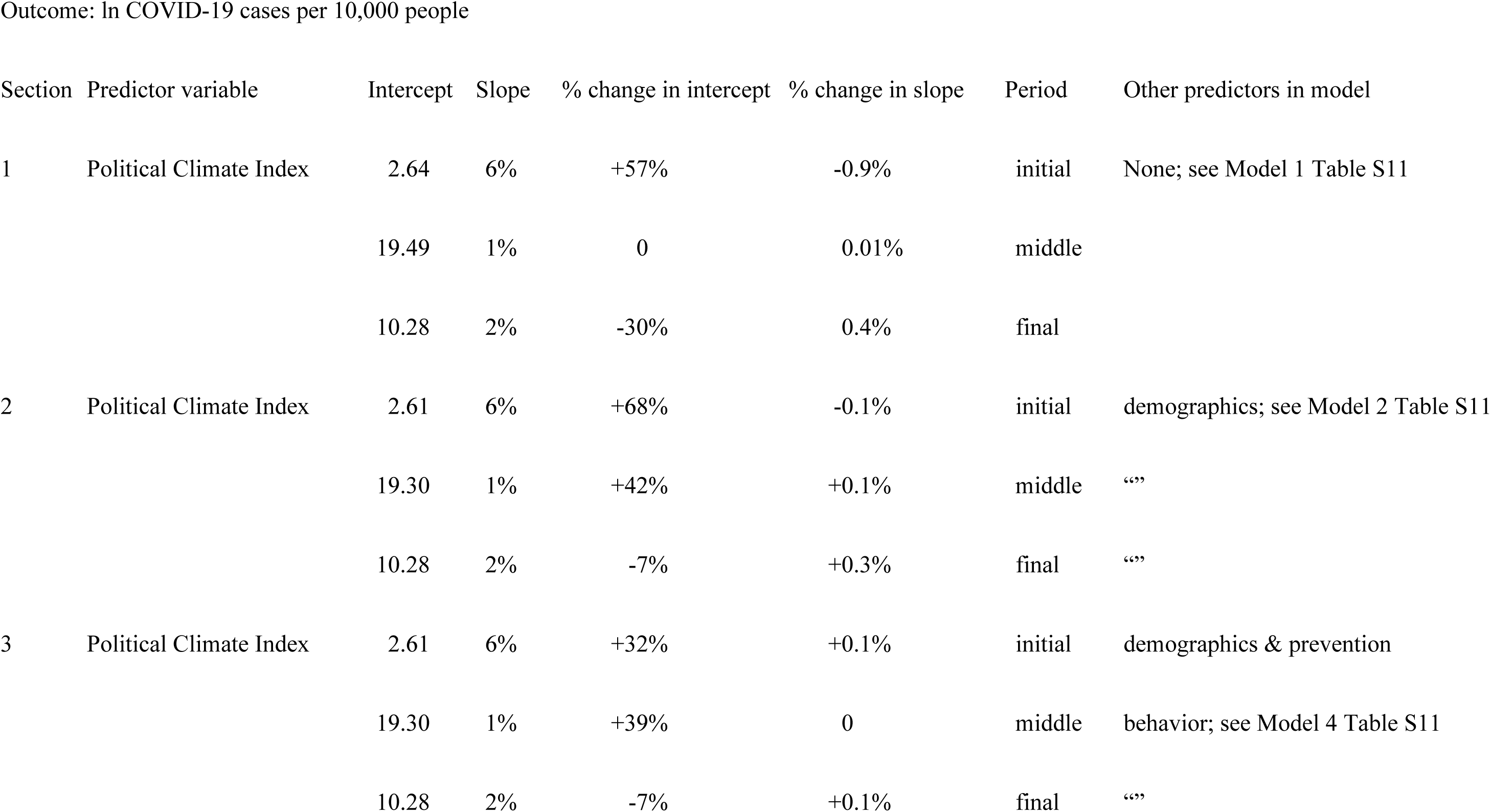
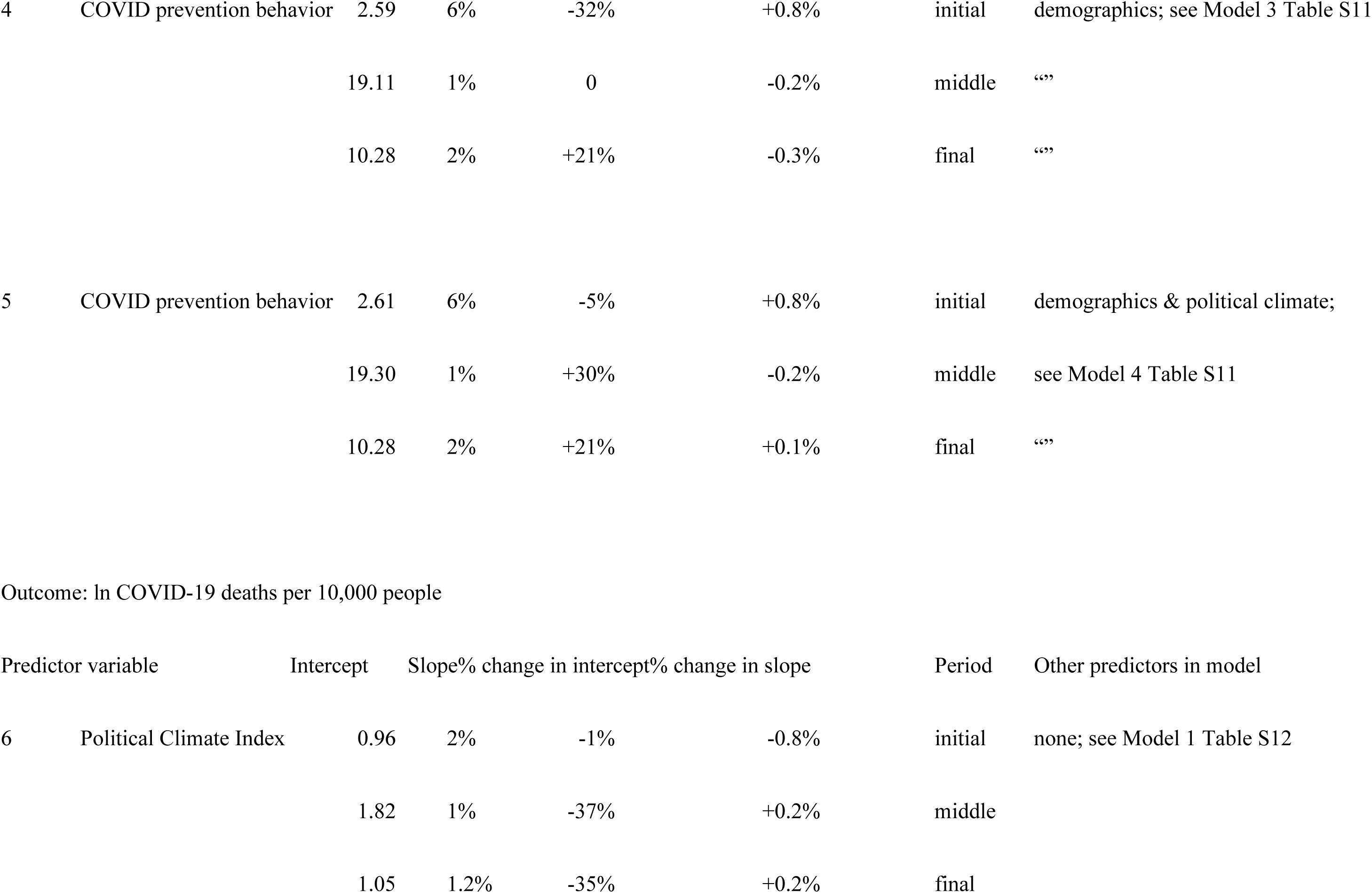
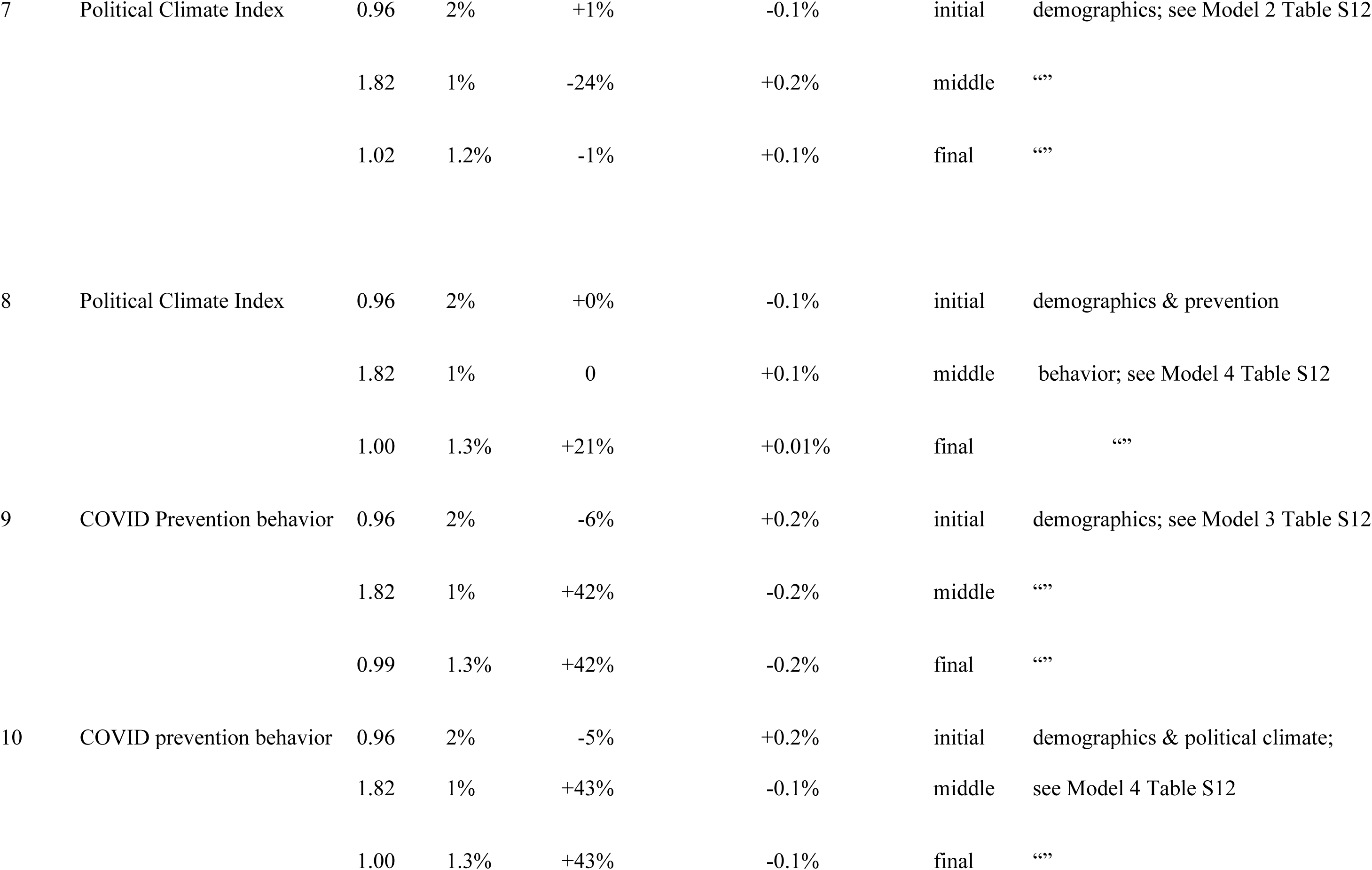
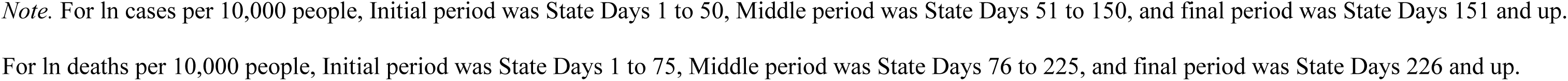
Intercepts, slopes, change in intercepts and change in slopes per unit of predictor variables by period, outcome (COVID-19 cases or deaths), and predictor set.

As predicted by Hypothesis 2, polities with more Republican-trusting political climates exhibited lower levels of adherence to COVID-19 prevention behaviors, *r* = -.67, *p* < .001 (see Table 3 for correlations of predictor variables across polities).

When we added preventative behavior as a predictor to the time models with demographics and political climate, we found that preventative behavior reduced the effects of political climate for cases and deaths (Sections 3 and 8 of Table 4). These results are consistent with Hypothesis 3 in that preventative behavior can be interpreted as partly mediating the association of political climate with both COVID-19 cases and deaths.

Given that political climate showed different patterns for cases and deaths, and that some of the health care access indicators correlated with political climate (see Table 3), we tested whether the relationship of cases to deaths was moderated by political climate. We found that it was. The average slope of .24 deaths to cases (in natural log per capita form) was reduced by .06 (25%) for every unit increase in Republican-trusting political climate (see Model 5 in Table S12). As Fig 3 shows, more Republican- trusting polities (shown in pink and blue) had a markedly weaker relationship between COVID-19 cases and deaths than did less Republican-trusting polities (shown in green and orange).

**Fig 3.**
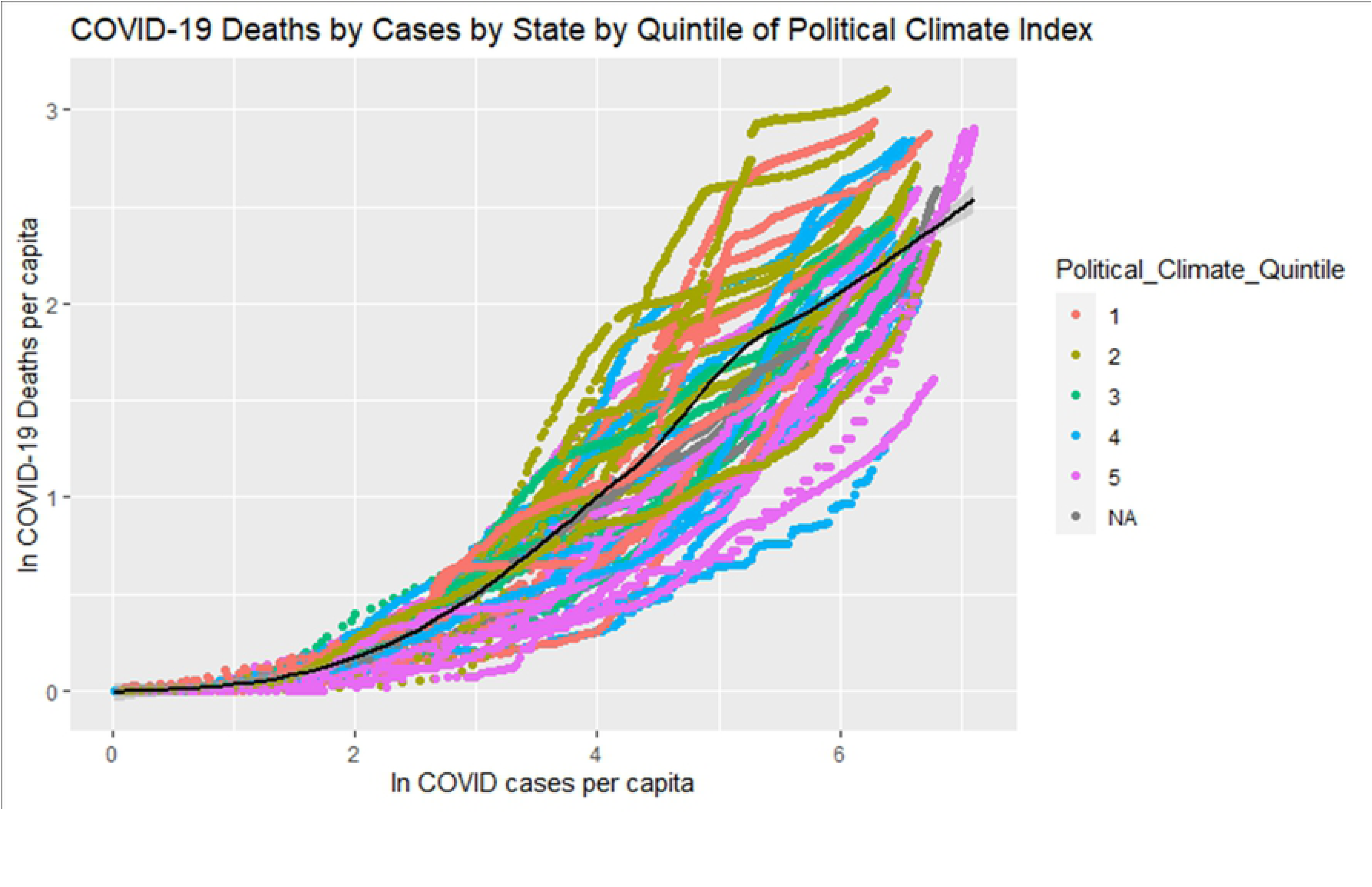
Relationship between COVID-19 cases and deaths as a function of political climate. Political climate quintiles from 1 least to 5 most, controlling for demographic effects. Black line illustrates average relationship between COVID cases and deaths.

### Health care access

Health care access influenced intercepts more strongly than it influenced daily slopes on COVID- 19 cases. For each unit increase in the latent medical access factor, case intercepts were 37% lower in the middle period and 27% lower in the final period, but were 3% higher in the initial period (see Model 1 in Table S13). For each unit increase in percent of people with no health insurance, the intercept for COVID- 19 cases was 84% higher in the initial period, 19% higher in the middle period, and 30% lower in the final period (see Model 2 in Table S13). COVID-19 case intercepts were 38% higher in the initial period, 25% higher in the middle period, and 13% lower in the final period for each unit increase in Z-scored ICU Beds per capita (see Model 3 in Table S13). These and other effects were highly similar when all health access predictor variables were included in the model (Model 4 in Table S13), and almost entirely accounted for political climate effects (see Table 4). Indeed, adding the political climate factor and its time parameter interactions to the combined health access model produced only one reliable (and small) political climate effect: the daily slope was higher by 0.2% for each unit of political climate index (see Model 5 in Table S13). For cases, then, Hypothesis 4 was confirmed; access to health care accounted for political climate effects on COVID-19 cases.

Access to health care also predicted intercepts for COVID-19 deaths. Latent medical access factor was unrelated to deaths in the middle period, but intercepts were 23% higher in the initial period and 21% lower in the final period for each unit of the medical care access index (Model 1 in Table S14). In the middle period, there were 27% fewer deaths for the intercept, with 3% increase in the initial period and 3% decrease in the intercept of the final period per unit of uninsured people (Model 2 in Table S14). Similarly, the middle period intercept was 16% lower, and in the initial period 1% lower, and in the final period 7% lower per unit of ICU Beds per capita (Model 3 in Table S14). Slopes were unrelated to access to health care; slopes changed by < |.04%| for all health care access predictors. Comparing the combined health access model (Model 4 in Table S14) with one that also included political climate index (Model 5 in Table S14) showed that there was little overlap in the effects of political climate and health care access. Political climate index effects remained reliable even with the three health care access measures and the larger model fit better, χ^2^ (6) = 274.22, *p* < 2 e^-16^. This implies that health care access did not account for all the political climate effects for COVID-19 deaths.

We tested whether adherence to COVID-prevention behaviors accounted for the effects of political climate on COVID-19 cases by adding the COVID-prevention behavior latent factor to the best models from Step 4. Models of cases in which both prevention behavior and political climate were entered showed that effects for both were weaker in the presence of the other than without it (compare Sections 3 to 2 and Sections 4 to 5 in Table 4).

Considering political climate effects and preventative behavior on COVID-19 cases together showed that some of the political climate effect could be attributed to polity differences in prevention behavior. The 68% increase in the initial intercept associated with each unit of political climate index decreased to 32% when preventative behavior was in the model (compare Models 2 and 4 in Table S11). The model considering political climate and demographics, χ^2^ (6) = 838, *p* < e^-16^, and the model considering preventative behavior and demographics, χ^2^ (6) = 537, *p* < e^-16^, fit worse than the model including all those predictors. These results are consistent with Hypothesis 3.

On COVID-19 deaths, political climate effects were largely accounted for by preventative behavior effects (compare Sections 7 & 8 in Table 4) although the reverse was not true (compare Sections 9 & 10 in Table 4). The model including both political climate and preventative behavior indices (and demographics; Model 5 in Table S12) fit better than the model including preventative behavior and demographics alone (Model 4 in Table S12), χ^2^ (5) = 114, *p* < e^-16^, and fit much better than the model including political climate and demographics alone (Model 2 in Table S12), χ^2^ (2) = 1084, *p* < e-^16^. These results for COVID-19 deaths are consistent with Hypothesis 3.

### COVID-preventative behavior

Consistent with smaller-scale studies of individuals [39], the more polities adhered to COVID-19 prevention behaviors, the fewer COVID-19 cases and deaths intercepts they had in the initial period, when prevention behavior was measured (see Table 5). Considering political climate effects and preventative behavior together as predictors showed that some of the political climate effect on cases and deaths could be attributed to polity differences in prevention behavior (see Sections 5 & 10 of Table 4), consistent with Hypothesis 3. As Fig 4 shows, polities with low prevention behavior transmitted COVID-19 more quickly.

**Fig 4.**
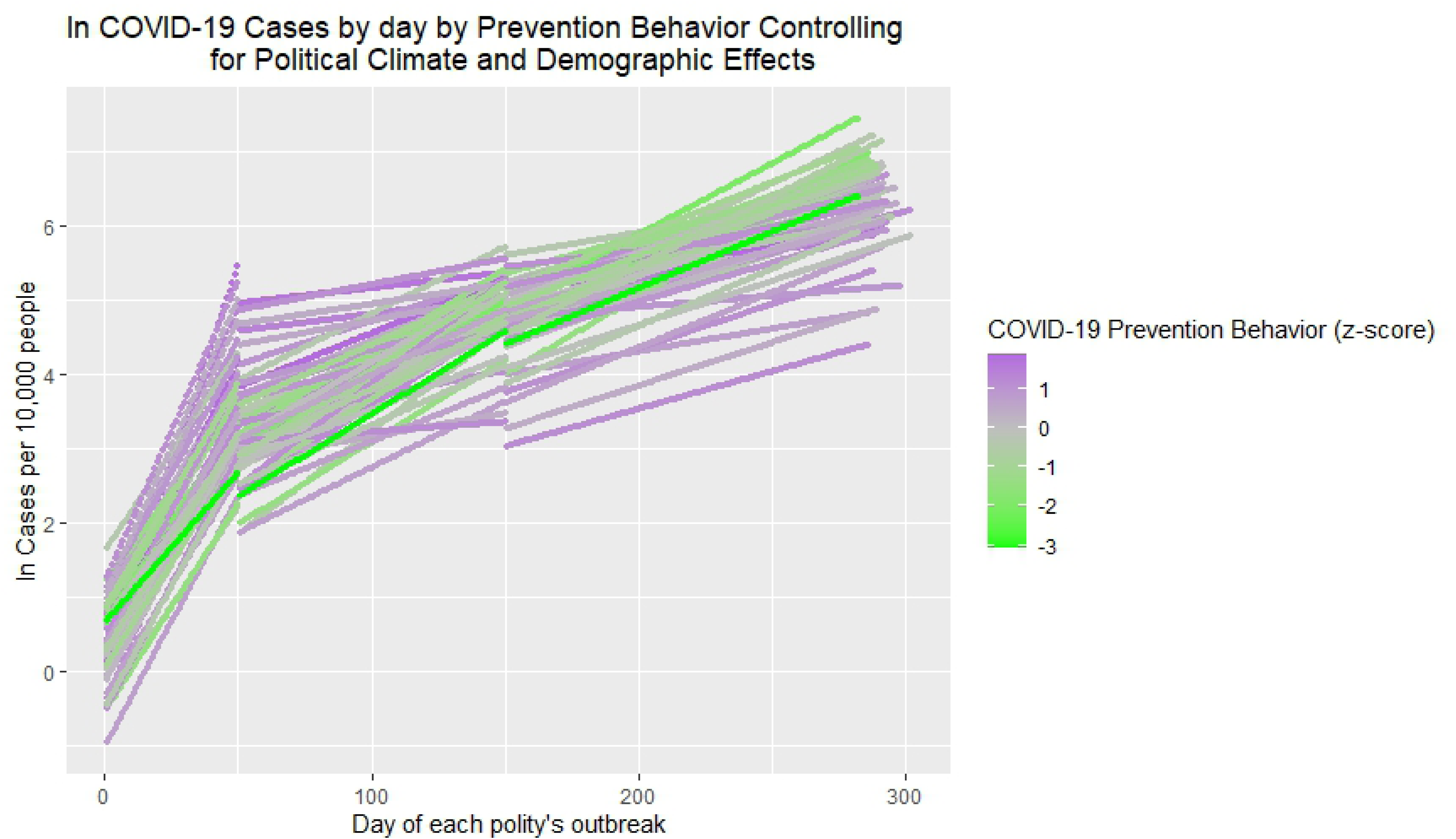
Model predicted natural log of COVID-19 cases per 10,000 people by day of state outbreak and prevention behavior factor Z-scored (“ZHEALTHBEHF”), controlling for demographic effects.

**Table 5.**
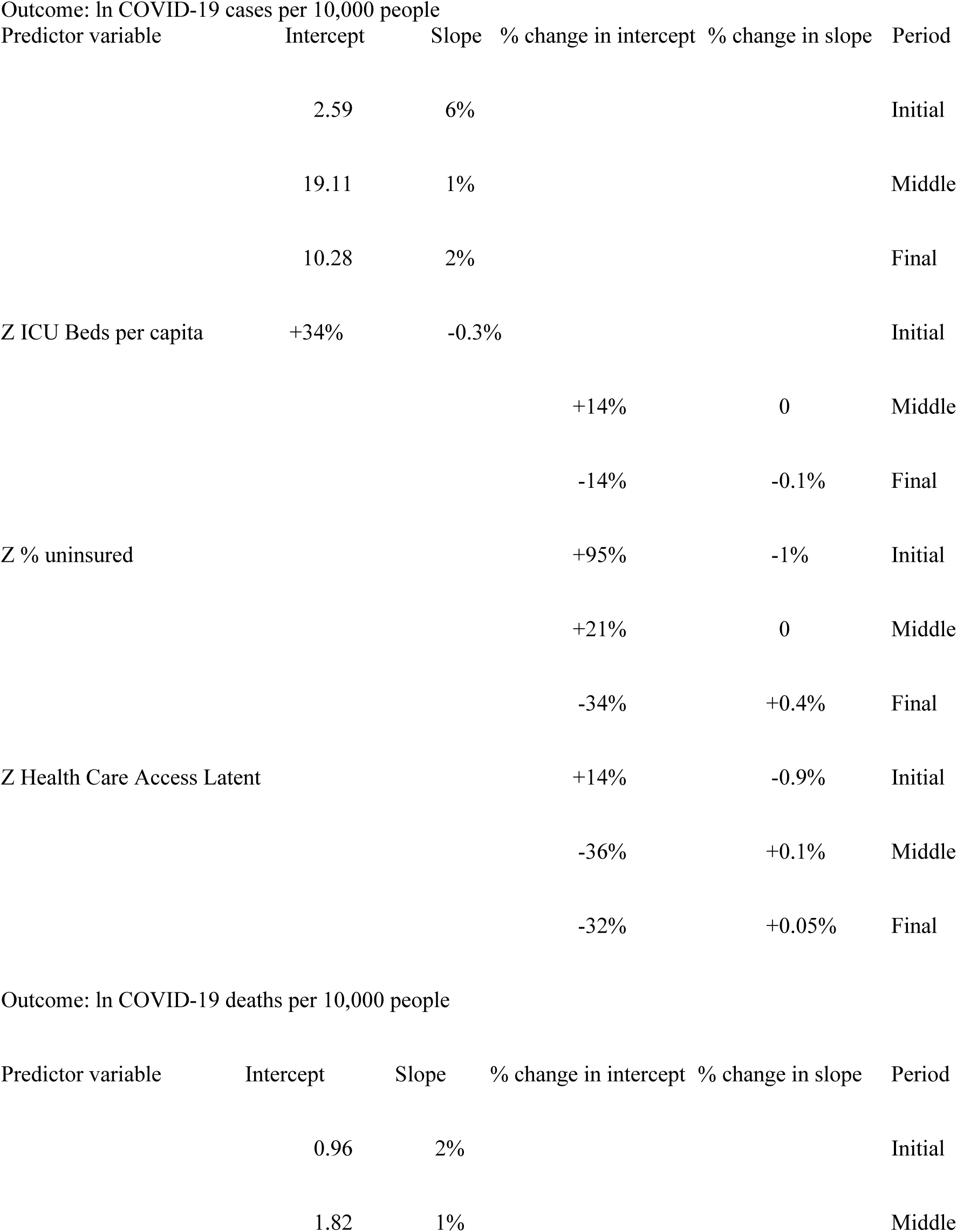
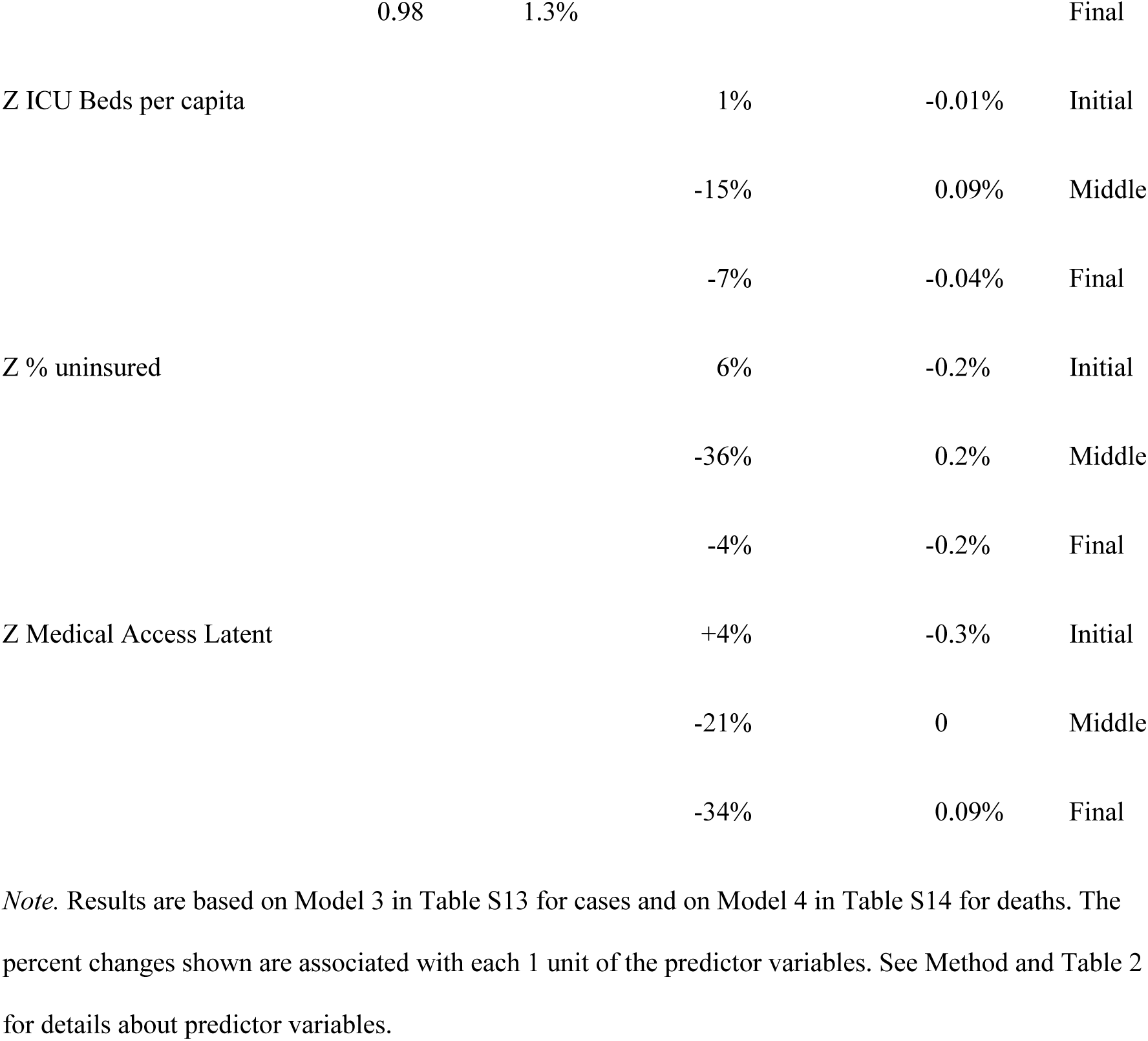
Intercepts, daily slopes, and changes due to health care access predictors for COVID-19 cases and deaths.

### Access to health care

Measures of access to health care included number of Intensive Care Unit (ICU) hospital beds per capita, percent of population without health insurance, and a latent medical access factor composed of per capita federally funded clinics, clinic visits, and provider sites (see Method and Table 2). We tested each health access measure separately and then combined reliable effects (see Tables S13, S14). The medical access predictors (see Table 3) and their effects (see Tables S13, S14) were largely independent.

Health care access measures influenced intercepts more strongly than they influenced daily slopes on COVID-19 cases (see Table S13) and deaths (see Table S14). Results suggested that access to health care improved COVID-19 outcomes (see Table 5). Polities with more uninsured people had substantially more COVID-19 cases in the initial and middle periods, whereas polities higher on latent medical access index had fewer cases in the middle and final periods. Deaths were less frequent in polities more health care access on the latent index and with more ICU Beds in the middle and final periods. Overall, then, better health care infrastructure was associated with fewer COVID-19 cases and deaths. These national results over the course of the year extend prior studies that used only part of the U.S., only part of the time period, and/or less extensive indicators of health care access than those we employed. [28, 44, 47].

## Discussion

The U.S. has the world’s top score on the Global Health Security Index, which is supposed to indicate health care infrastructure readiness, and yet the U.S. has the most COVID-19 deaths; evidently, political capacity is critical for responding to pandemics [40, see 41, 42, 43 for international comparisons of COVID-19 outcomes]. Our study helps to explain why. We found that Republican-trusting political climates had elevated COVID-19 risks due to faster COVID-19 transmission, less preventative behavior, poorer healthcare infrastructure and lower access to health insurance. Our results extend shorter-term studies associating communities’ Republican voting, affiliation, and/or leadership with worse COVID-19 outcomes [44, 35]. Our study further found evidence for two proximal vectors of this political climate risk: low access to health care and low compliance with recommended COVID-19 prevention behaviors.

### Health Care Access

It was only 20 days from the first to the last state outbreaks, but the fact that outbreaks that started days later had fewer deaths per capita suggests that medical and nursing personnel figured out more effective means to treat acute COVID-19 even within the first few weeks. For example, in April, 2020, staff treating patients suffering from hypoxemia realized that patients fared better when placed in prone positions [45, 46].

The meaning of health care access effects on COVID-19 cases and deaths depends on the direction of the effects. In some studies, shelter-in-place orders are positively related to case and death counts [47], presumably because such orders are a *response* to increased disease incidence. Whereas we found evidence that access to health care improved COVID-19 outcomes and that lack of health insurance was associated with more cases and deaths, we also found evidence that access to health care might enable detection of cases and treatment. In particular, more cases were detected in the initial and middle periods in polities with more ICU Beds per capita, and the latent score on medical access was associated with higher intercepts for cases and deaths in the initial period. Similarly, a county-level study of US COVID-19 cases through May 25, 2020 found that more primary care physicians per capita was positively associated with case counts.

[45] Like [45], who argue that ICU beds are an indicator of the “health system’s ability to handle critically ill patients,” we found that more per capita ICU beds was negatively related to COVID-19 deaths in the middle and final periods (see Table 5).

To our knowledge, prior research to the present study has not uncovered the fact that there is a weaker relationship between COVID-19 cases and deaths in more Republican-trusting political climates. Generally, death counts are assumed to be more accurate than case counts [45]. Our results suggest that greater health care access is associated with more cases due to ability to avail oneself of health care facilities. Coupled with the finding of lower correspondence between cases and deaths even controlling for time changes and demographics, this interpretation has important implications for public health officials and politicians who decide on acute health policy because it implies that death-counts in polities with less health care infrastructure may be misleadingly low.

### Preventative Behavior

Our results showed that part of the effects of political climate could be attributed to political- climate-related differences in adherence to COVID-19 prevention behaviors such as avoiding crowds and wearing masks. In particular, preventative behavior measured in the initial period of outbreaks was negatively related to COVID-19 cases and deaths in that period. Short-term studies conducted on individuals [37] and on smaller polities have found that preventative behavioral measures do reduce transmission rates [48, 49]. In 2021, a new important preventative behavior became getting vaccinated against SARS-CoV-2. Corroborating our theoretical analysis of political climate, an August, 2021 national study of US counties shows that vaccination and death rates correspond to political climate, as indicated by the 2016 election vote spread for Trump-Biden [50]. Thus, the influence of Republican-trusting political climate, with its minimization of public health practices and of the dangers of COVID-19 has continued even with the advent of effective vaccines.

### Learning from Experience

Studying the pandemic over time leads to consideration of what the public and others learn from it. We found that cases and deaths were especially associated with greater urbanity in the initial and middle periods. This may have led people in rural areas to assume that COVID-19 would not affect them; there are significant differences in levels of trust in government and also in access to health care between urban and rural US [32]. Unfortunately, anyone who inferred that COVID-19 was a coastal, city problem made an erroneous assumption. By October 1, 2021, rural areas had high vaccination refusal rates, and high rates of a much more contagious and equally lethal COVID-19 mutation (delta) that was overwhelming their health-care facilities [51].

Conversely, polities with more preventative behavior in the initial period had higher case and death counts in the last period. If such polities dropped their previous effective habits because their communities had seen early outbreaks wane, they became vulnerable to COVID-19 transmission again by doing so. A basic psychophysical principle is that people notice change when it is large compared to a base rate, such as in the initial growth period of contagious disease spread. In fact, a study showed that governors made lock-down decisions in accordance with this principle [52]. Models and experience demonstrate that change following a substantial increase in COVID-19 transmission is too late to prevent steep increases of this highly contagious disease.

### Caveats

The present study could not be experimental, and so it lacks the possibility of strong causal inference. Observational studies leave open the possibility of unexamined causal variables. Our study did, however, use reliable data sources encompassing all the states and DC for 300 days, omitting only US territories because predictor data for them were not available. The causality of political climate as a significant factor in COVID-19 spread is plausible for several reasons. First, it is parsimonious, theoretically-grounded, and consistent with a number of studies that used smaller polities (e.g., counties), or were shorter-term, or smaller-scale [32, 42, 45]. Second, the present study found evidence for two plausible mediators of the influence of Republican-trusting political climate on COVID-19 cases and deaths: limited access to health care (assessed in multiple ways) and low public cooperation with COVID-19 prevention behaviors. Although the polity unit of analysis used was somewhat appropriate because state governors often did set COVID-19 policies, it is also somewhat crude both for assessing climate and policies.

Stronger results might have obtained with a finer-grained unit of locale for assessing political climate, but the consistency of the present results with studies using a variety of other methods, from anonymous cell phone searches [33, 34] to opinion polls [26, 48] to medical records [42, 45], lends credibility to the findings. The reliably weaker relationship of the rate of cases to deaths for polities higher on the political climate index bears further investigation because it may imply that standards for detecting and reporting cases and deaths are not uniform. Alternatively, inequality in health care access may not only endanger the health of people living in underserved areas, but may threaten the integrity of health data, which may lead to misinformed and poorer policy-making. Systematic investigation of this finding and its implications for policies and practices is warranted.

### Summary

Trust in others and in institutions is necessary to invite the large-scale cooperation among members of the public with various institutions necessary to mount an effective response to a novel pandemic. Variability around the globe [45, 46] as well as variability across the U.S. [39] demonstrates that such trust is supported more in some political climates than in others. Our conceptualization of political climate need not identify particular ideologies nor particular parties, let alone official political Parties. But it happens that the U.S. has a political Party that is identifiable in its ideological promotion of mistrust of science and government and in curtailing public health programs. We predicted and found that polities who elected more Republicans to state (or in DC’s case, city) legislatures also had publics who trusted President Trump and the White House more to handle COVID-19, and those same polities had more uninsured residents, were more likely not to have expanded Medicare, and were less likely to have engaged in mask wearing, avoiding crowds, and avoiding close contact as means of preventing COVID-19 spread. Critically, results showed that lack of health insurance was strongly related to more COVID-19 cases in the initial period, whereas more prevention behavior in the initial period was strongly related to fewer COVID-19 cases and deaths in the initial period. Both the policies (limited access to health care) and ideology (COVID-19 public health skepticism) touted by Republican Party officials were followed by residents, and the more they did so, the more risk they took with their own health and lives and those of their neighbors.

## Data Availability

The data files are available from the Open Science Framework website https://osf.io/hwgzy/?view_only=922c0cf9225847c3907e5b0b7db7416a.

## Acknowledgments

No funding supported this research. The authors declare no conflicts of interest. The authors thank Chloe Lafosse for research assistance, Dr. Emil Coman for comments on a draft, and Dr. Timothy Moore from UConn’s Statistical Consulting Services for help with figures.

## Footnotes

1 χ^2^(1) = 5.56, *p* < .02

2 *SD* = 2.7% and *SD* = 2.2% respectively, partial *η^2^* = .40.

3 χ^2^(1) = 16.73, *p* < .001.

4 Tables and Figures beginning with “S” are in Supplementary Materials.

